# Prediction of Diabetes in Middle-Aged Adults: A Machine Learning Approach

**DOI:** 10.1101/2023.11.26.23299025

**Authors:** Gideon Addo, Bismark Amponsah Yeboah, Michael Obuobi, Raphael Doh-Nani, Seidu Mohammed, David Kojo Amakye

**Affiliations:** Department of Statistics and Actuarial Science, College of Science, Kwame Nkrumah University of Science and Technology, Kumasi, Ghana; Department of Pathology, School of Medicine and Dentistry, Kwame Nkrumah University of Science and Technology, Kumasi, Ghana; Department of Mathematical Sciences, College of Science, University of Texas at El Paso, Texas, United States of America

**Keywords:** Diabetes Prediction, Middle-Aged Adults, Diabetes Symptoms, Machine Learning Models, Predictive Performance Metrics

## Abstract

**Background:** Diabetes is a serious and progressive medical condition demanding efficient diagnostic methods, especially since its associated symptoms overlap with the symptoms of other medical conditions. While various studies have explored early detection of diabetes across different age groups, there is a notable gap in specific attention to middle-aged adults. This study explicitly focused on this demographic, aiming to assess associations between symptoms and diabetes status, investigate the relevance and relative influence of certain symptomatic and demographic features in the prediction of diabetes, and identify the most efficient machine learning (ML) model for predicting diabetes.

**Methods:** Utilizing a dataset from a previous study conducted in the Sylhet Diabetes Hospital in Bangladesh, India, comprising 520 participants, including both diabetic and non-diabetic patients, we extracted and analyzed demographic and symptom-related information from 296 middle-aged adults aged from 40 to 60 years. Employing chi-square tests, we evaluated symptom-diabetes associations, while utilizing the Boruta algorithm to investigate symptom importance and influence. Seven ML models namely, K-Nearest Neighbor (KNN), Naïve Bayes (NB) classifier, Support Vector Machines with linear, polynomial, and radial basis function kernels, Random Forest (RF) classifier, and Logistic Regression were then assessed for optimal predictive performance.

**Results:** Out of the 296 participants of this study, 179 (60%) were diabetic. Significant associations were found between diabetes status in middle-aged adults and symptoms such as polyuria, polydipsia, weakness, sudden weight loss, partial paresis, polyphagia, and visual blurring, as confirmed by the p-values of their respective chi-square tests. All features studied, including demographics and symptoms, were confirmed as relevant for predicting diabetes in middle-aged adults. Notably, polyuria, polydipsia, gender, alopecia, irritability, and sudden weight loss were identified as the most influential features. Among the seven ML models, RF showed the highest sensitivity (98.59%), while KNN excelled in specificity (97.83%). RF demonstrated the best accuracy (96.58%) and area under the curve score (96.00%), making it the most efficient ML model for predicting diabetes among middle-aged adults.

**Conclusion:** The findings of this study emphasize the importance of using diabetes-related symptoms for early detection of diabetes within the middle-aged adult population. The RF model demonstrated robust diagnostic capabilities, emphasizing its potential in predicting diabetes in middle-aged adults. Further exploration of genetic, lifestyle, and environmental factors is warranted to enhance the understanding and diagnostic accuracy in this demographic.

## INTRODUCTION

Diabetes stands as a persistent medical condition marked by heightened blood glucose levels and disruptions in the metabolism of fats and proteins [1]. This chronic ailment encompasses a cluster of symptoms linked to hyperglycemia, signifying elevated sugar levels in the bloodstream [2]. The intricate process by which the body converts food into energy is altered in diabetes. Typically, the body breaks down food into sugar and glucose, releasing them into the bloodstream. Elevated blood sugar levels prompt the pancreas to release insulin, acting as a key to facilitate the entry of sugar into the body’s cells for energy utilization [3]. In diabetes, either insufficient insulin is produced or the body fails to use insulin effectively, leading to excessive blood sugar lingering in the bloodstream. Over time, this condition can give rise to severe health complications such as heart disease, vision impairment, and kidney disorders [3]. The global prevalence of diabetes has surged, quadrupling since 1980, with an estimated 422 million people worldwide affected, according to the World Health Organization [4].

The Centers for Disease Control and Prevention (CDC) identifies three primary types of diabetes: Type 1 (T1D), Type 2 (T2D), and Gestational (GD) diabetes. T1D is a heterogeneous disorder characterized by the destruction of pancreatic beta cells, resulting in absolute insulin deficiency [5]. As an autoimmune disorder, T1D is considered the most common and severe among autoimmune diseases [6], though it accounts for only 5-10% of global diabetes cases [7,8]. T1D can manifest at any age, with the highest incidence observed in children [9]. On the other hand, T2D is a progressive disorder marked by deficiencies in insulin secretion and increased insulin resistance, leading to abnormal glucose metabolism and related metabolic disruptions [10]. Representing approximately 90% of all diabetes cases, T2D predominantly affects middle-aged adults [11]. Symptoms of T2D may develop gradually over several years, sometimes going unnoticed for an extended period [12]. Lastly, GD is a less common form of diabetes occurring in pregnant women without pre-existing diabetes, leading to elevated blood sugar levels during pregnancy. The outcomes of GD include an elevated risk of maternal cardiovascular disease and type 2 diabetes [13]. Additionally, infants born to mothers with GD may experience macrosomia (excessive birth weight) and complications during birth [13]. Globally, 14% of all pregnancies are affected by GD [14].

Irrespective of the diabetes type, neglecting early signs and symptoms can result in severe long-term complications, including cardiovascular issues, kidney problems, vision impairment, and neuropathic conditions. Recognizable early signs that necessitate diabetes diagnosis and investigation encompass frequent urination (polyuria), excessive thirst (polydipsia), sudden weight loss, excessive hunger (polyphagia), blurred vision, unexpected weakness, dry skin, delayed healing, and frequent infections [15]. Given the overlap of symptoms with other medical conditions, there is a crucial need for highly efficient and user-friendly diagnostic methods to aid healthcare professionals and patients in determining the diabetes status of individuals presenting with similar symptoms.

In light of the escalating global incidence and mortality rates of diabetes [16], numerous studies have delved into understanding the etiology, diagnosis, and management of the condition. For the effective management of diabetes early detection is crucial, as the condition can deteriorate over time if not properly addressed or remains undetected. While many diagnostic studies rely on laboratory-based tests, some are outdated [17,18,19], despite some having high accuracy rates [19]. The recent surge in Artificial Intelligence (AI) has prompted a growing number of studies utilizing Machine Learning (ML) techniques to detect and screen diabetes across various demographics and populations, including gender [20–22], income classes [23], and age groups such as infants [24], children and teenagers [25–27]. Surprisingly, few, if any, of these studies explicitly concentrate on employing ML techniques to screen and diagnose diabetes in middle-aged adults. This study aims to fill this gap by focusing on using ML methods to detect diabetes specifically in the middle-aged adult population.

The primary objectives of this study are to: 1) assess associations between symptoms displayed by middle-aged adults and their diabetes status, 2) investigate the relevance and relative influence of some demographic and symptomatic features in predicting diabetes status among middle-aged adults, and 3) identify the most effective ML model for predicting diabetes status in middle-aged adults. The significance of this study lies in its potential to provide valuable insights into the associations between symptoms, demographic factors, and diabetes status in the middle-aged population. By achieving its objectives to assess these associations and identify the most effective ML model, the study aims to contribute crucial information to clinical practice. The findings may offer a refined and efficient approach to diagnosing and treating diabetes in middle-aged adults, ultimately improving healthcare outcomes in this specific demographic.

## MATERIALS AND METHODS

### Study Design and Data Source

This research utilized the dataset derived from a previous study conducted by Islam et al. [28] in India, where they aimed at constructing a predictive tool for early-stage diabetes risks predictions. The dataset, publicly available on Mendeley Data [29], include information on patient demographics and diabetes-related symptoms obtained from 520 participants through a questionnaire. These participants were patients visiting the Sylhet Diabetes Hospital in Bangladesh, and included both diabetic and non-diabetic individuals showing signs similar to that of diabetes. The dataset contains comprehensive information on patient symptoms, demographics, and diabetes status, making it suitable for addressing the objectives of this current investigation.

For our study, we specifically extracted data for 296 middle-aged adults within the original dataset, where the inclusion criterion was for individuals aged between 40 and 60 years. Exclusion criterion was for all other patients who fell outside this age bracket.

### Features and Feature Selection

Within the dataset employed for this study, 17 variables were identified, with 16 serving as features and one as the outcome variable. The outcome variable contained information about a patient’s diabetes status, while the features included details on patient demographics and symptoms linked to diabetes, as outlined in Table 1. All features, excluding the “Age” variable, were nominal with two classes. Notably, our dataset contained no missing values.

**Table 1.**
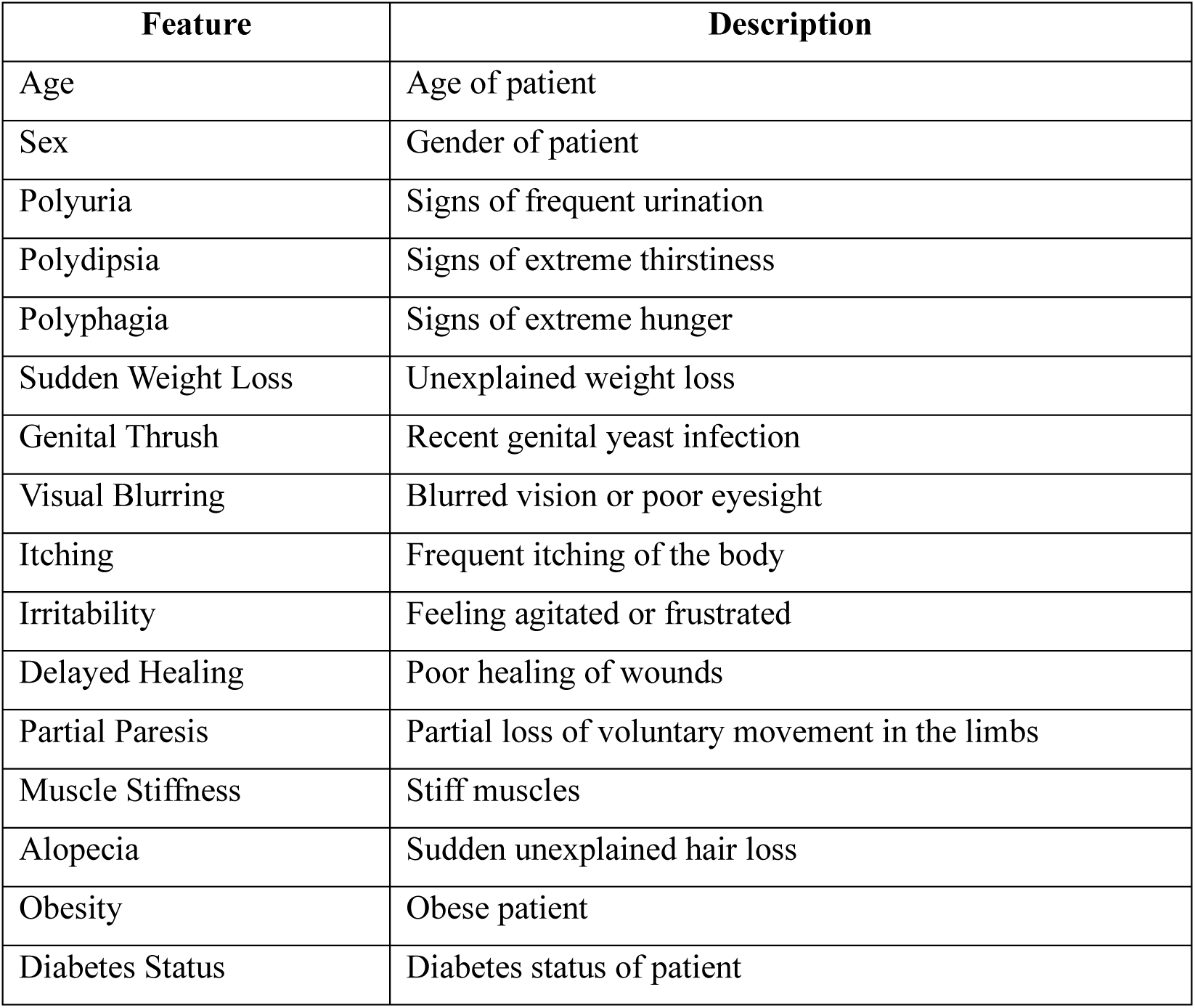
Description of features used in this study.

To assess the influence of features on predicting diabetes status, we employed the Boruta algorithm, accessible through the Boruta package in R [30], for feature selection. The Boruta algorithm, recognized for its all-relevant approach, utilizes a wrapper algorithm to identify influential features. Leveraging the random forest algorithm, it conducts a comprehensive search for relevant features by comparing original attribute importance with estimates derived from permuted copies, progressively eliminating irrelevant features. This algorithm is adaptable, accommodating any classification method outputting variable importance.

For a visual assessment of feature influence and relevance, we utilized a Boruta algorithm-generated plot, illustrated in Figure 1. In this plot, the vertical axis represents feature importance, while the horizontal axis represents features. The color of each boxplot indicates the relevance of the feature in predicting the outcome variable. Blue boxplots correspond to the minimal, average, and maximum Z score of a shadow feature, while red, green, and yellow boxplots represent Z scores of rejected, confirmed, and tentative features, respectively. Confirmed features are those recognized as significant predictors of the outcome variable. Rejected features, on the other hand, are deemed non-important in predicting the outcome variable. Tentative features are predictors for which the Boruta algorithm could not conclusively determine their importance.

**Figure 1.**
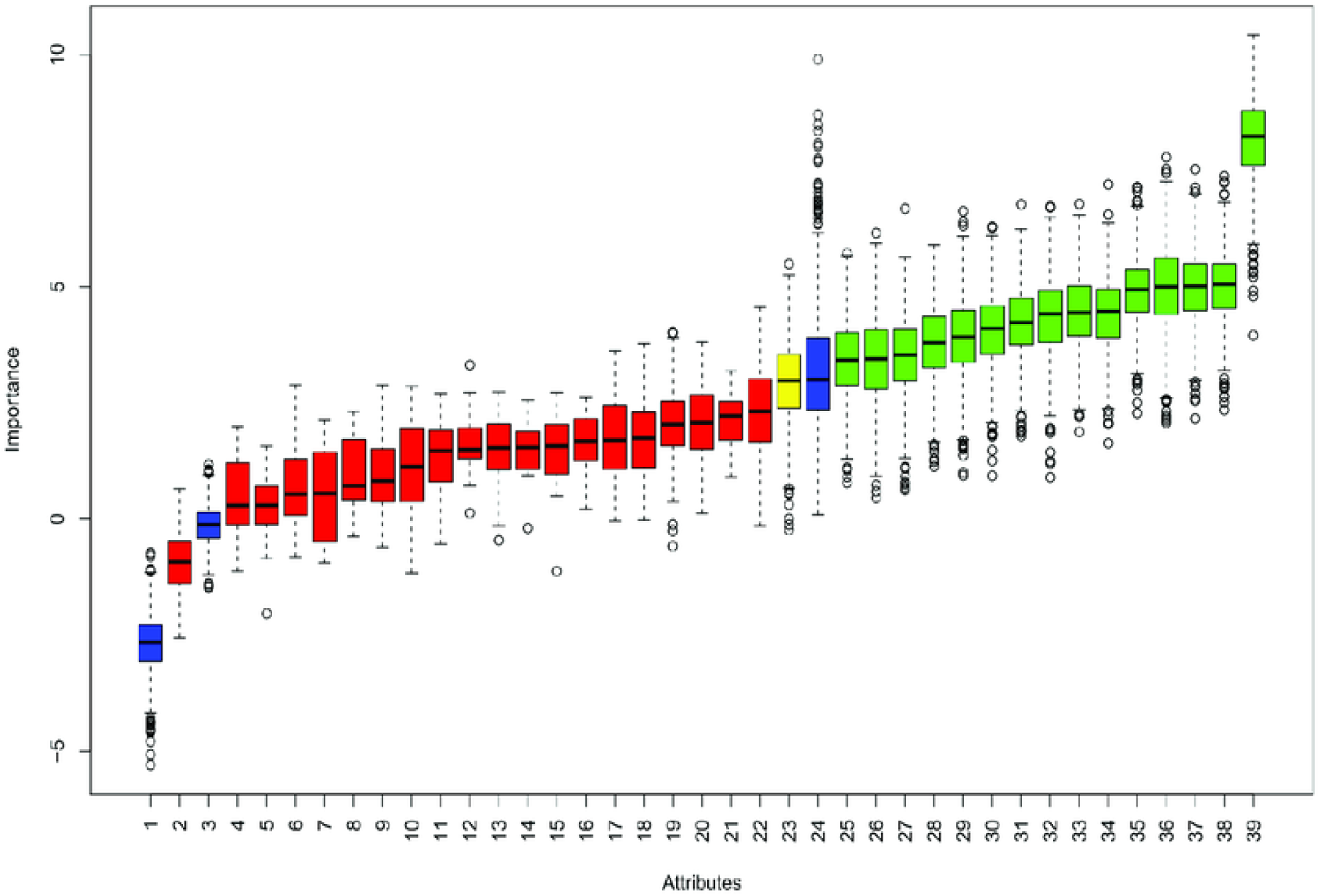
Illustration of a sample Boruta algorithm generated feature importance plot.

### Statistical Analysis

To prepare our data for analysis, we conducted data preprocessing. Except for the “Age” variable, all other variables were nominal with two classes. Utilizing one-hot encoding, we encoded all nominal variables, representing the presence of a category with “1” and its absence with “0”. Additionally, we applied the min-max scaling technique to standardize the “Age” variable’s scale, bringing its values within the range of 0 to 1.

In this study, descriptive statistics were employed to characterize the primary outcome variable and all other predictor variables. Frequencies and percentages were used to describe the nominal variables. To examine associations between symptoms exhibited by middle-aged adults and their diabetes status, we employed the chi-square test of association. This statistical method evaluates the dependence or independence between two nominal variables, determining whether a significant association exists based on observed and expected frequencies within a contingency table.

Furthermore, to determine the most efficient ML algorithm for predicting diabetes within the middle-aged adult population, we assessed the performance of seven distinct algorithms. These algorithms include K-Nearest Neighbor (KNN), Naïve Bayes (NB) classifier, Support Vector Machines (SVM) with linear, polynomial, and radial basis function (RBF) kernels, Random Forest (RF) classifier, and Logistic Regression (LR). In evaluating the performance of the ML models, various performance metrics were utilized, including Accuracy, Sensitivity, Specificity, Positive Predictive Value (PPV), Negative Predictive Value (NPV), and Receiver Operating Characteristic (ROC) curve analysis with their associated Area Under the Curve (AUC) values.

Sensitivity measures a model’s ability to correctly identify positive cases, highlighting its effectiveness in capturing true positives. Specificity gauges a model’s accuracy in identifying negative cases, emphasizing its proficiency in avoiding false positives. PPV assesses the likelihood that a positive prediction is accurate, offering insights into the reliability of positive results. NPV estimates the probability of a negative prediction being correct, indicating the reliability of negative outcomes. Accuracy quantifies the overall correctness of a model’s predictions, providing a comprehensive measure of its performance. AUC evaluates a model’s ability to distinguish between different classes, offering a concise summary of its discriminative power across various thresholds. In this study, the selection of the optimal ML model for predicting diabetes status among middle-aged adults was based on the one demonstrating both the highest accuracy and AUC scores.

The chosen level of significance throughout the study was set at 0.05. All statistical analyses were carried out using the R programming language (version 4. 3. 2).

## RESULTS

### Assessing the associations between symptoms displayed by middle-aged adults and their diabetes status

Table 2 displays symptoms in middle-aged patients, both with and without diabetes, along with the p-values from chi-square tests assessing associations with diabetes status. Among the 296 participants, 179 (60%) were diabetic. Symptoms like polyuria, polydipsia, weakness, sudden weight loss, partial paresis, polyphagia, and visual blurring are more prevalent in diabetic middle-aged patients, and rare in their non-diabetic counterparts. This emphasizes the significance of these symptoms as potential indicators of diabetes in this age group. Timely interventions should be considered when these symptoms manifest, enhancing healthcare providers’ vigilance in detecting diabetes in middle-aged individuals.

**Table 2.**
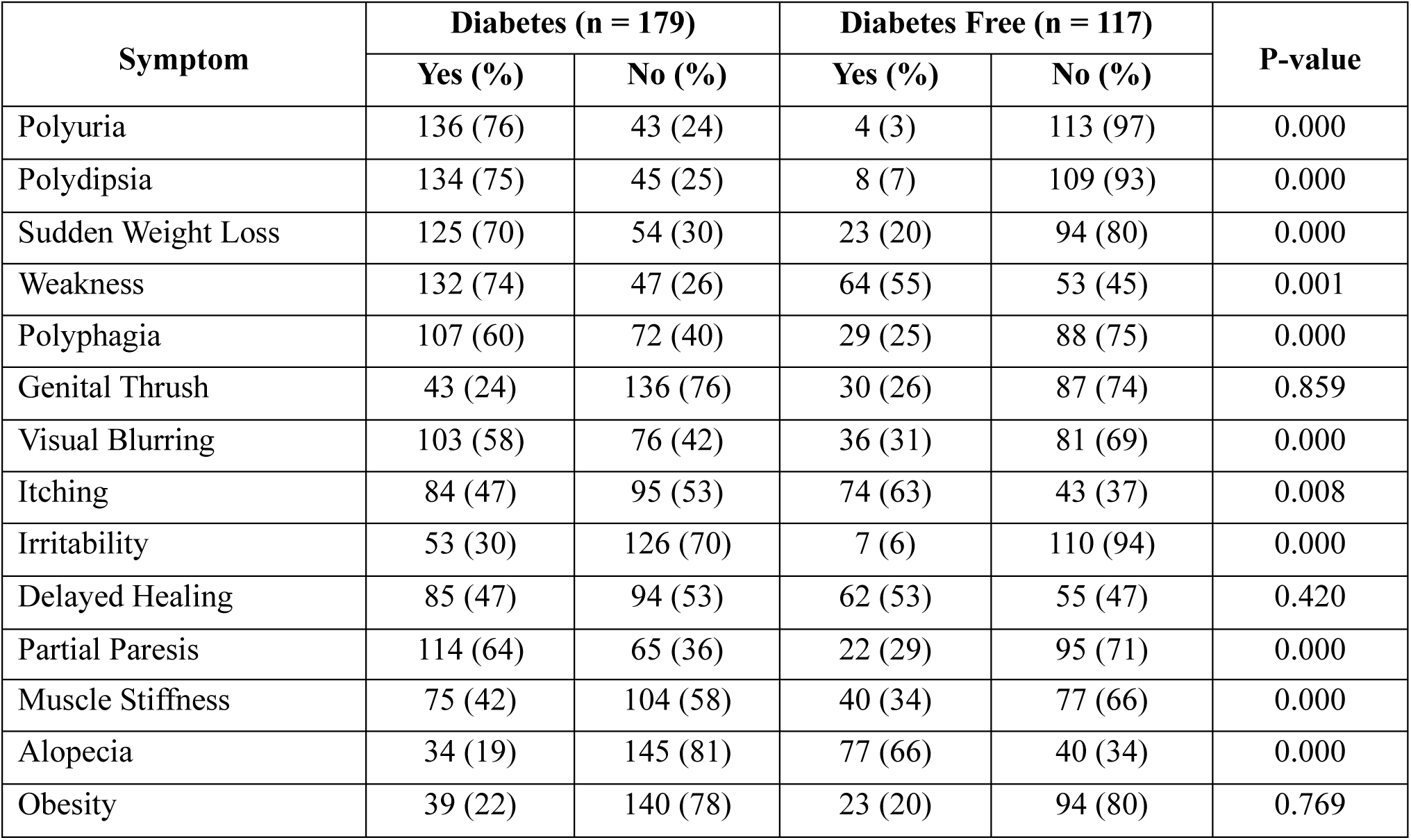
Results of the chi-square tests between diabetes status and symptoms.

| Symptom | Diabetes (n = 179) |  | Diabetes Free (n = 117) |  | P-value |
| --- | --- | --- | --- | --- | --- |
|  | Yes (%) | No (%) | Yes (%) | No (%) |  |
| Polyuria | 136 (76) | 43 (24) | 4 (3) | 113 (97) | 0.000 |
| Polydipsia | 134 (75) | 45 (25) | 8 (7) | 109 (93) | 0.000 |
| Sudden Weight Loss | 125 (70) | 54 (30) | 23 (20) | 94 (80) | 0.000 |
| Weakness | 132 (74) | 47 (26) | 64 (55) | 53 (45) | 0.001 |
| Polyphagia | 107 (60) | 72 (40) | 29 (25) | 88 (75) | 0.000 |
| Genital Thrush | 43 (24) | 136 (76) | 30 (26) | 87 (74) | 0.859 |
| Visual Blurring | 103 (58) | 76 (42) | 36 (31) | 81 (69) | 0.000 |
| Itching | 84 (47) | 95 (53) | 74 (63) | 43 (37) | 0.008 |
| Irritability | 53 (30) | 126 (70) | 7 (6) | 110 (94) | 0.000 |
| Delayed Healing | 85 (47) | 94 (53) | 62 (53) | 55 (47) | 0.420 |
| Partial Paresis | 114 (64) | 65 (36) | 22 (29) | 95 (71) | 0.000 |
| Muscle Stiffness | 75 (42) | 104 (58) | 40 (34) | 77 (66) | 0.000 |
| Alopecia | 34 (19) | 145 (81) | 77 (66) | 40 (34) | 0.000 |
| Obesity | 39 (22) | 140 (78) | 23 (20) | 94 (80) | 0.769 |

The p-values from chi-square tests highlight strong associations between diabetes status and symptoms like polyuria, polydipsia, weakness, sudden weight loss, partial paresis, polyphagia, and visual blurring in middle-aged individuals. These results emphasize the importance of these symptoms as strong indicators of diabetes in this age group. In contrast, symptoms such as obesity, delayed healing, and genital thrush show weaker associations with diabetes status in middle-aged individuals, suggesting that these conditions may not be as specific to diabetes in this age group. Healthcare providers should consider additional diagnostic criteria alongside these symptoms.

### Investigating the relevance and relative influence of the demographic and symptomatic features in the prediction of diabetes status

Figure 2 visually presents the relevance and relative importance of each variable employed in our study to predict diabetes status in middle-aged patients. The plot reveals clinical and statistical implications that can enhance the understanding and application of these symptoms in the diagnosis of diabetes in this specific population.

**Figure 2.**
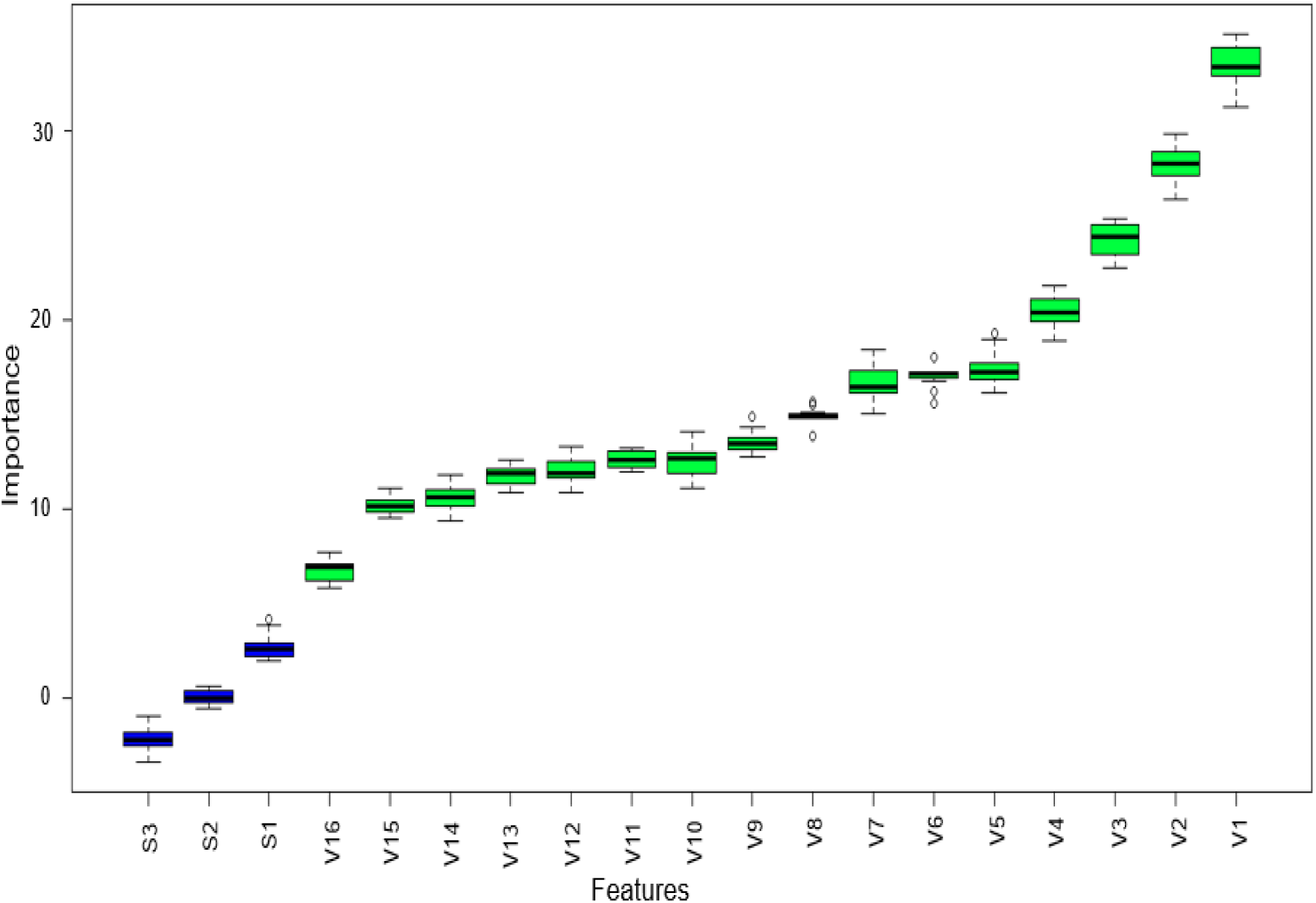
Relative importance of features in predicting diabetes status among middle-aged adults. *V1 - Polyuria; V2 - Polydipsia; V3 - Gender; V4 - Alopecia; V5 - Irritability; V6 - Sudden Weight Loss; V7 - Age; V8 - Partial Paresis; V9 - Polyphagia; V10 - Delayed Healing; V11 - Visual Blurring; V12 - Muscle Stiffness; V13 - Weakness; V14 - Itching; V15 - Obesity; V16 - Genital Thrush; S1 - ShadowMax; S2 - ShadowMean; S3 – ShadowMin.*

From the plot, we observe that certain predictors, namely polyuria, polydipsia, gender, alopecia, irritability, and sudden weight loss, are highly influential features in descending order of importance when it comes to predicting the diabetes status of middle-aged patients. Healthcare professionals should pay close attention to these predictors, as they can serve as strong indicators of diabetes in middle-aged individuals, potentially expediting the diagnostic process. Conversely, features such as genital thrush, itching, obesity, and muscle stiffness are shown to be less important when predicting diabetes status in middle-aged patients. This statistical insight suggests that while these features still play a role in diabetes diagnosis, their relative importance is lower. Statistically, the green-colored boxplots associated with all features signify that the Boruta algorithm identified each variable as an important contributor to predicting diabetes status in middle-aged patients.

### Identifying the most effective ML model for predicting diabetes in middle-aged adults

Table 3 presents a comprehensive evaluation of the performance of the seven ML models used in predicting diabetes in our study, considering their sensitivity, specificity, PPV, NPV, and accuracy scores. From the table, the RF model demonstrated superior sensitivity at 98.59%, outperforming other models, while KNN excelled in specificity with a score of 97.83%. The KNN model achieved the highest PPV at 98.39%, indicating its reliability in identifying diabetes, while the RF model led in NPV at 97.73%, indicating its reliability in identifying non-diabetes. Overall accuracy favored the RF model with a score of 96.58%, emphasizing its superiority in predicting both diabetic and non-diabetic cases among middle-aged adults. These findings offer valuable insights for healthcare practitioners when selecting a ML model for diabetes prediction in this demographic and highlight the importance of tailoring approaches based on model strengths and weaknesses.

**Table 3.** Performance metrics associated with each machine learning model.

| Model | Sensitivity (%) | Specificity (%) | PPV (%) | NPV (%) | Accuracy (%) |
| --- | --- | --- | --- | --- | --- |
| KNN (k = 5) | 85.92 | 97.83 | 98.39 | 81.82 | 90.60 |
| NB | 97.18 | 78.26 | 87.34 | 94.74 | 89.74 |
| SVM (Linear) | 91.55 | 95.65 | 97.01 | 88.00 | 93.16 |
| SVM (Polynomial) | 92.96 | 95.65 | 97.06 | 89.80 | 94.02 |
| SVM (RBF) | 91.55 | 93.48 | 95.59 | 87.76 | 92.31 |
| RF | 98.59 | 93.48 | 95.89 | 97.73 | 96.58 |
| LR | 92.96 | 95.65 | 97.06 | 89.80 | 94.02 |

### ROC and AUC Analysis

Figure 3 illustrates the ROC curves and their corresponding AUC values for each model used in predicting diabetes among middle-aged adults in this study. AUC values are crucial indicators of a model’s ability to distinguish between positive and negative cases. From the plot, the RF model stood out with an impressive AUC of 96.0%, surpassing other models. While AUC values were closely aligned, SVM (Polynomial) and LR had overlapping curves with identical AUC values of 94.3%. In summary, the RF model exhibited superior discriminative performance, making it the optimal choice for predicting diabetes in middle-aged adults. The consistently high AUC scores of the models underscore their robust predictive capabilities.

**Figure 3.**
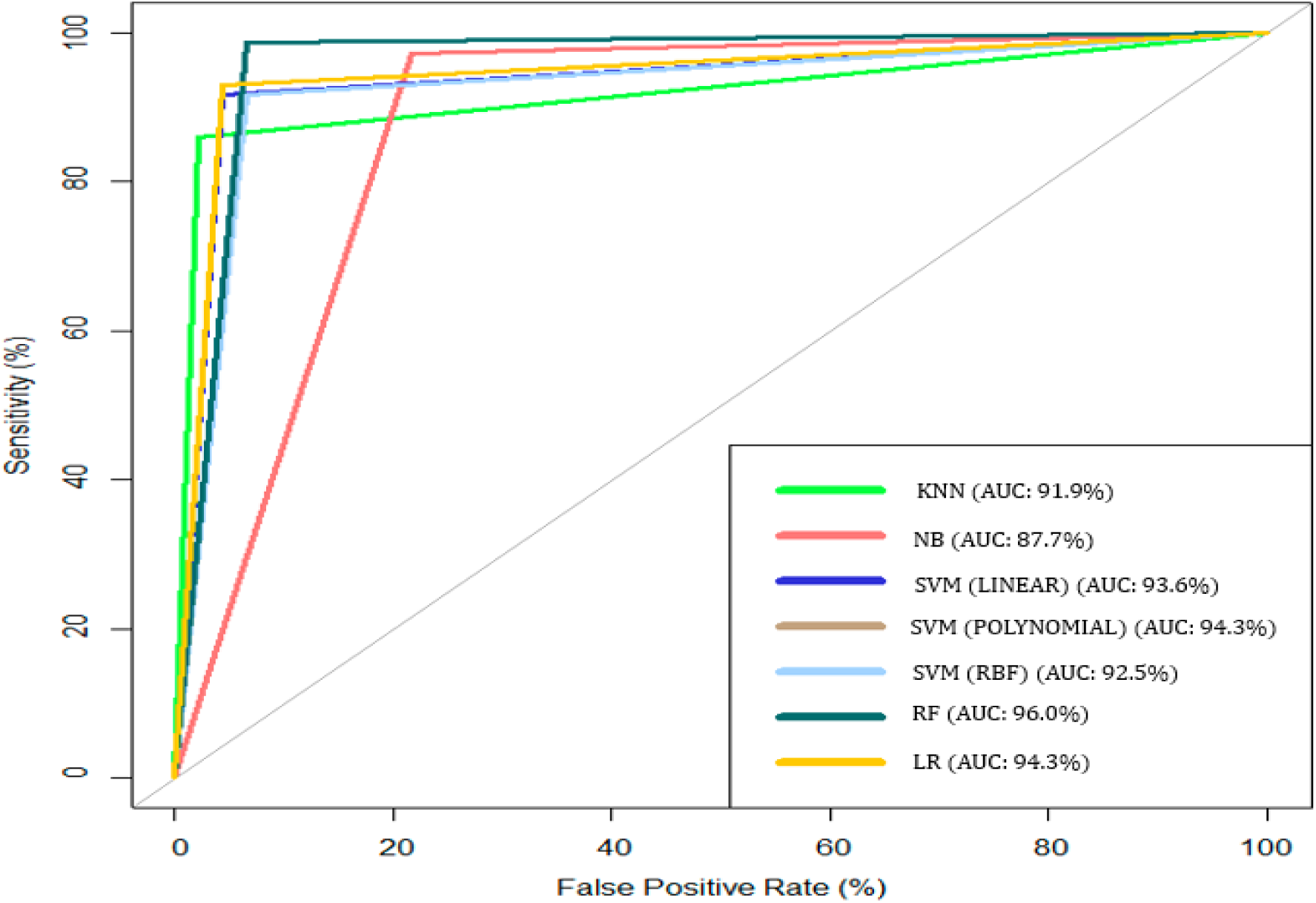
ROC curves with their corresponding AUC scores of the machine learning models of this study.

## DISCUSSION

Diabetes is a significant health concern, and its symptoms can remain subtle or unnoticed for extended periods, particularly in T2D [16,31]. Since key diabetes symptoms may resemble those of other medical conditions, individuals often prioritize addressing those conditions. Consequently, diabetes diagnosis tends to occur in later disease stages when symptoms become more severe. This study explored the associations between diabetes status and specific symptoms in middle-aged adults, including both diabetic and non-diabetic individuals. Our findings indicate that among middle-aged adults, the prevalent symptoms associated with diabetes include polyuria, polydipsia, weakness, sudden weight loss, polyphagia, partial paresis, and visual blurring. Conversely, these symptoms are less common among their non-diabetic counterparts.

Several studies across different populations have reported findings consistent with ours [31–34]. Devi et al. [31], in their investigation into the prevalence of T2D among the Meiteis of Manipur in India, identified similar symptoms among T2D patients, including polyuria, polyphagia, polydipsia, fatigue, and weight loss, all of which were significantly associated with diabetes. Similarly, Kumar et al. [32] observed a parallel discovery in a different age demographic, noting that children with diabetes commonly exhibited symptoms such as polyuria, polydipsia, weight loss, and fatigue. In another study, Pawar et al. [33] found a strong indication of diabetes in patients displaying polydipsia, polyuria, polyphagia, or a combination of these symptoms. The consistency of our study’s findings with those of previous studies in different populations suggests that middle-aged adults exhibit symptoms of diabetes similar to those in other demographics, with the classical symptoms being polyuria, polyphagia, polydipsia, visual blurring, and fatigue.

In contrast, our study revealed that symptoms such as obesity, delayed healing, and genital thrush were not linked to the diabetes status of middle-aged adults. This contradicts previous research that established a strong association between T2D and obesity [35–37]. For instance, a 2011 study by Barness [37] found a significant increase in the risk of diabetes among overweight and obese women, with overweight women facing a 28-fold risk and obese women an astonishing 93-fold risk compared to normal-weight women. The reasons for these discrepancies between our study’s findings and existing research are not entirely clear. One plausible explanation is that a majority of our study participants may be suffering from T1D, which is typically not associated with obesity. However, it’s noteworthy that Pawar et al. [33] also reported similar findings to ours.

Moreover, our study found no association between delayed wound healing and diabetes among middle-aged adults, contradicting previous research [38,39]. Fahey et al. [38] concluded in their study that delayed healing in diabetes is linked to altered leucocyte infiltration and wound fluid IL-6 levels during the late inflammatory phase of the wound healing process. Our findings suggest that certain symptoms traditionally associated with diabetes may not always be indicative in this specific demographic. Therefore, further research is warranted to explore the factors influencing these differences and refine our understanding of diabetes manifestation in middle-aged adults.

Additionally, we explored the relevance and relative influence of symptoms and demographic factors in predicting diabetes in middle-aged adults. Our study’s findings revealed a hierarchy of influential symptoms, with polyuria, polydipsia, alopecia, and irritability being the most critical indicators of diabetes among middle-aged individuals. Gender, specifically being female, also played a noteworthy role in this age group. This hierarchy provides valuable insights for healthcare professionals, emphasizing the crucial relevance of these symptoms in diagnosing diabetes among middle-aged adults. Notably, polyuria and polydipsia emerged as robust early signals for diabetes in this specific population. Conversely, symptoms such as itching, obesity, and muscle stiffness, although identified as important determinants in predicting diabetes among middle-aged adults, had relatively low influence in predicting diabetes within this age demographic. Healthcare professionals can use this information to optimize diagnostic assessments, potentially expediting the identification of diabetes in middle-aged adults.

Precise classification of diabetes status holds paramount importance, as it prevents the progression of the condition and facilitates potential reversals, especially in cases of prediabetes. The recent surge in AI has spurred numerous research studies, aiming to streamline the identification and diagnosis of diabetes, resulting in the need for models with exceptionally high accuracy measures. Several studies employing ML techniques for diabetes predictions consistently yield classification models with outstanding performance scores [40–46].

In our study, all the ML models exhibited commendable sensitivity scores, signifying their ability to accurately predict diabetes cases among the middle-aged adult population. Notably, the RF model stood out with the highest sensitivity score of 98.59%. The specificity scores were generally high and closely aligned, with the KNN algorithm emerging as the top performer in predicting non-diabetes cases among middle-aged adults, boasting a specificity score of 97.83%. Notably, the RF model excelled in overall accuracy and AUC, achieving scores of 96.58% and 96.00%, respectively. This aligns with the consensus in previous studies, highlighting the effectiveness of ensemble ML techniques in predicting or diagnosing diabetes across diverse populations [42,44,47,48,49].

Healthcare practitioners are encouraged to leverage these findings when choosing ML models for diabetes prediction within the middle-aged adult population, emphasizing the potential benefits of ensemble learning techniques, particularly the RF model, in achieving high accuracy and reliability in diabetes classification.

### Limitations

In conducting this research, it is essential to acknowledge certain limitations that may influence the breadth and applicability of our findings. Firstly, our study relied on a dataset from a prior investigation in India, which could potentially constrain the representativeness of the data to the broader middle-aged adult population. Regional variations in healthcare systems and lifestyle factors could impact the generalizability of the results to other settings. Secondly, a notable limitation is the assumption in our research that all types of diabetes share the symptoms employed in our study. In reality, the symptoms chosen may not universally apply to all diabetes types, and the clinical implications of the predictive model may be specific to the prevalent diabetes types in our dataset. Additionally, our study’s exclusive focus on particular symptoms for diabetes prediction may overlook other pertinent symptoms or risk factors that contribute to a comprehensive understanding of diabetes in middle-aged adults. The analysis may not have included additional symptoms that could be relevant. Moreover, it is crucial to recognize that our study did not control for potential confounding variables such as genetic factors, lifestyle factors, medication usage, comorbid conditions, socioeconomic status, and the geographic location of the study’s participants. The presence of these uncontrolled variables could introduce variability in our findings, potentially impacting the generalizability of our results.

## CONCLUSION

In conclusion, the findings of this study shed light on the associations between symptoms and the diabetes status of middle-aged adults. The identification of specific symptoms, including polyuria, polydipsia, weakness, sudden weight loss, partial paresis, polyphagia, visual blurring, irritability, itching, muscle stiffness, and alopecia, as associated with diabetes status provides valuable insights for both clinicians and individuals in this demographic. Notably, symptoms such as obesity, delayed healing, and genital thrush were not found to be associated with diabetes within this age group. This warrants the need for future research given that these symptoms have been established by previous research as associated symptoms of diabetes in other demographics.

Furthermore, the study highlights the varying degrees of importance among the identified features in predicting diabetes status. Polyuria, polydipsia, gender, alopecia, irritability, and sudden weight loss emerged as the most influential factors in predicting diabetes among middle-aged adults. On the other hand, symptoms like genital thrush, obesity, and itching were comparatively less influential in determining diabetes status within this demographic.

The RF classifier’s robust discriminative capabilities in predicting diabetes status indicates the importance of employing advanced ML techniques for accurate and reliable predictions. However, while this study provides important insights, there remains a need for further research to explore additional factors and potential interactions that may influence diabetes status among middle-aged adults. Future studies could delve into the role of genetic predispositions, lifestyle factors, and environmental influences to enhance our understanding and improve the precision of predictive models. Such endeavors are essential for the development of targeted interventions and personalized healthcare strategies, ultimately contributing to better outcomes for individuals in this age group at risk of diabetes.

## Data Accessibility

The findings of this study are supported by data available under the Creative Commons Attribution 4.0 International License (CC BY 4.0). The dataset can be accessed via the Mendeley Data [29], offering unrestricted entry to the compiled data for reuse and redistribution. Researchers and stakeholders are welcome to explore and employ the dataset, adhering to the stipulations of the Creative Commons license.

## Data Availability

All data produced are available online at https://doi.org/10.17632/7zcc8v6hvp.1

## REFERENCES

1. Roglić G. WHO Global report on diabetes: A summary. International Journal of Noncommunicable Diseases. 2016; 1(1): 3. Available from: 10.4103/2468-8827.184853

2. Chou CY, Hsu D, Chou CH. Predicting the Onset of Diabetes with Machine Learning Methods. Journal of Personalized Medicine. 2023; 13(3): 406. Available from: 10.3390/jpm13030406

3. What is Diabetes? Centers for Disease Control and Prevention. 2023. Available from: https://www.cdc.gov/diabetes/basics/diabetes.html

4. World Health Organization: WHO. Diabetes. 2021. Available from: https://www.who.int/news-room/facts-in-pictures/detail/diabetes

5. Maahs DM, West NA, Lawrence JM, Mayer-Davis EJ. Epidemiology of Type 1 diabetes. Endocrinology and Metabolism Clinics of North America. 2010; 39(3): 481–97. Available from: 10.1016/j.ecl.2010.05.011

6. Dariya B, Chalikonda G, Srivani G, Alam A, Nagaraju GP. Pathophysiology, etiology, epidemiology of Type 1 diabetes and computational approaches for immune targets and therapy. Critical Reviews in Immunology. 2019; 39(4): 239–65. Available from: 10.1615/critrevimmunol.2019033126

7. American Diabetes Association. Diagnosis and classification of diabetes mellitus. Diabetes Care. 2010; 33 Supplement 1: S62–9. Available from: 10.2337/dc09-s062

8. Mobasseri M, Shirmohammadi M, Amiri T, Vahed N, Fard HH, Ghojazadeh M. Prevalence and incidence of type 1 diabetes in the world: a systematic review and meta-analysis. Health Promotion Perspectives. 2020; 10(2): 98–115. Available from: 10.34172/hpp.2020.18

9. Gale EAM. The rise of childhood Type 1 diabetes in the 20th century. Diabetes. 2002; 51(12): 3353–61. Available from: 10.2337/diabetes.51.12.3353

10. Meigs JB, Muller DC, Nathan DM, Blake DR, Andres R. The natural history of progression from normal glucose tolerance to Type 2 diabetes in the Baltimore Longitudinal Study of Aging. Diabetes. 2003; 52(6): 1475–84. Available from: 10.2337/diabetes.52.6.1475

11. Zeng B, Lu Y, Hajifathalian K, Bentham J, Di Cesare M, Danaei G, et al. Worldwide trends in diabetes since 1980: a pooled analysis of 751 population-based studies with 4·4 million participants. The Lancet. 2016; 387(10027): 1513–30. Available from: 10.1016/s0140-6736(16)00618-8

12. Ramachandran A. Know the signs and symptoms of diabetes. The Indian Journal of Medical Research. 2014; 140(5): 579.

13. Plows JF, Stanley JC, Baker PN, Reynolds CM, Vickers MH. The Pathophysiology of Gestational diabetes mellitus. International Journal of Molecular Sciences. 2018; 19(11): 3342. Available from: 10.3390/ijms19113342

14. Juan J, Yang H. Prevalence, prevention, and lifestyle intervention of gestational diabetes mellitus in China. International Journal of Environmental Research and Public Health. 2020; 17(24): 9517. Available from: 10.3390/ijerph17249517

15. Blair ME. Diabetes mellitus review. Urologic Nursing. 2016; 36(1): 27. Available from: 10.7257/1053-816×.2016.36.1.27

16. World Health Organization: WHO. Diabetes. 2023. Available from: https://www.who.int/news-room/fact-sheets/detail/diabetes

17. Emancipator K. Laboratory diagnosis and monitoring of diabetes mellitus. American Journal of Clinical Pathology. 1999; 112(5): 665–74. Available from: 10.1093/ajcp/112.5.665

18. Sacks DB, Bruns DE, Goldstein DE, Maclaren NK, McDonald JM, Parrott M. Guidelines and recommendations for laboratory analysis in the diagnosis and management of diabetes mellitus. Clinical Chemistry. 2002; 48(3): 436–72. Available from: 10.1093/clinchem/48.3.436

19. Kaur G, Lakshmi PVM, Rastogi A, Bhansali A, Jain S, Teerawattananon Y, et al. Diagnostic accuracy of tests for type 2 diabetes and prediabetes: A systematic review and meta-analysis. PLOS ONE. 2020; 15(11): e0242415. Available from: 10.1371/journal.pone.0242415

20. Man B, Schwartz A, Xia Y, Gerber BS. Individualized Diabetes Risk Prediction in Women with a History of Gestational Diabetes. Diabetes. 2018; 67 Supplement 1. Available from: 10.2337/db18-1293-p

21. Li T, Quan H, Zhang H, Lin L, Lü L, Ou Q, et al. Type 2 diabetes is more predictable in women than men by multiple anthropometric and biochemical measures. Scientific Reports. 2021; 11(1). Available from: 10.1038/s41598-021-85581-z

22. Zhang X, Zhao X, Huo L, Yuan N, Sun J, Du J, et al. Risk prediction model of gestational diabetes mellitus based on nomogram in a Chinese population cohort study. Scientific Reports. 2020; 10(1). Available from: 10.1038/s41598-020-78164-x

23. Boutilier JJ, Chan TCY, Ranjan M, Deo S. Risk stratification for early detection of diabetes and hypertension in Resource-Limited Settings: Machine Learning analysis. Journal of Medical Internet Research. 2021; 23(1): e20123. Available from: 10.2196/20123

24. Fernández-Edreira D, Liñares-Blanco J, Fernández-Lozano C. Machine Learning analysis of the human infant gut microbiome identifies influential species in type 1 diabetes. Expert Systems With Applications. 2021; 185: 115648. Available from: 10.1016/j.eswa.2021.115648

25. Kushwaha S, Srivastava RN, Jain R, Sagar V, Aggarwal AK, Bhadada SK, et al. Harnessing machine learning models for non-invasive pre-diabetes screening in children and adolescents. Computer Methods and Programs in Biomedicine. 2022; 226: 107180. Available from: 10.1016/j.cmpb.2022.107180

26. Hu H, Lai T, Farid F. Feasibility study of constructing a screening tool for adolescent diabetes detection applying machine learning methods. Sensors. 2022; 22(16): 6155. Available from: 10.3390/s22166155

27. Diabetes Prediction in Teenagers using Machine Learning Algorithms. IEEE Conference Publication | IEEE Xplore. 2023. Available from: https://ieeexplore.ieee.org/abstract/document/10112286

28. Islam MdM, Ferdousi R, Rahman S, Bushra HY. Likelihood prediction of diabetes at early stage using data mining techniques. In: Advances in intelligent systems and computing. 2019. p. 113–25. Available from: 10.1007/978-981-13-8798-2_12

29. Joseph LP. Diabetes datasets. Mendeley Data. Aug 15; Available from: 10.17632/7zcc8v6hvp.1

30. Kursa MB, Rudnicki WR. Feature Selection with theBorutaPackage. Journal of Statistical Software. 2010; 36(11). Available from: 10.18637/jss.v036.i11

31. Devi KBL, Meitei KT, Singh SD. Prevalence of Type 2 diabetes and its signs and symptoms among the Meiteis of Manipur, India. Journal of the Anthropological Survey of India. 2022; 72(1): 59–70. Available from: 10.1177/2277436×221136955

32. Kumar A, Kaplowitz PB. Patient age, race and the type of diabetes have an impact on the presenting symptoms, latency before diagnosis and laboratory abnormalities at time of diagnosis of diabetes mellitus in children. Journal of Clinical Research in Pediatric Endocrinology. 2009; 1(5): 227–32. Available from: 10.4274/jcrpe.v1i5.227

33. Pawar SD, Thakur P, Radhe B, Jadhav H, Behere V, Pagar V. The accuracy of polyuria, polydipsia, polyphagia, and Indian Diabetes Risk Score in adults screened for diabetes mellitus type-II. Medical Journal of Dr DY Patil University. 2017; 10(3): 263. Available from: 10.4103/0975-2870.206569

34. Pawar SD, Naik JD, Prabhu PM, Jatti GM, Jadhav S, Radhe B. Comparative evaluation of Indian Diabetes Risk Score and Finnish Diabetes Risk Score for predicting risk of diabetes mellitus type II: A teaching hospital-based survey in Maharashtra. Journal of Family Medicine and Primary Care. 2017; 6(1): 120. Available from: 10.4103/2249-4863.214957

35. Boles A, Kandimalla R, Reddy PH. Dynamics of diabetes and obesity: Epidemiological perspective. Biochimica Et Biophysica Acta (BBA) - Molecular Basis of Disease. 2017; 1863(5): 1026–36. Available from: 10.1016/j.bbadis.2017.01.016

36. Mokdad AH, Ford ES, Bowman BA, Dietz WH, Vinicor F, Bales VS, et al. Prevalence of Obesity, Diabetes, and Obesity-Related Health Risk Factors, 2001. JAMA. 2003; 289(1): 76. Available from: 10.1001/jama.289.1.76

37. Barnes AS. The epidemic of obesity and diabetes: trends and treatments. Texas Heart Institute Journal. 2011; 38(2): 142–4.

38. Fahey TJ, Sadaty A, Jones WG, Barber A, Smoller BR, Shires GT. Diabetes impairs the late inflammatory response to wound healing. Journal of Surgical Research. 1991; 50(4): 308–13. Available from: 10.1016/0022-4804(91)90196-s

39. Tang Y, Zhang MJ, Hellmann J, Kosuri M, Bhatnagar A, Spite M. Proresolution therapy for the treatment of delayed healing of diabetic wounds. Diabetes. 2013; 62(2): 618–27. Available from: 10.2337/db12-0684

40. Kumari S, Kumar D, Mittal M. An ensemble approach for classification and prediction of diabetes mellitus using soft voting classifier. International Journal of Cognitive Computing in Engineering. 2021; 2: 40–6. Available from: 10.1016/j.ijcce.2021.01.001

41. Viswanatha V, Ac R, Murthy D, Thanishka. Diabetes Prediction using Machine learning approach. Social Science Research Network. 2023; Available from: 10.2139/ssrn.4533862

42. Sadhu A, Jadli A. Early-stage diabetes risk prediction: A comparative analysis of classification algorithms. International Advanced Research Journal in Science, Engineering and Technology (IARJSET). 2021; 8(2): 193–201.

43. Xue J, Min F, Ma F. Research on Diabetes Prediction Method based on Machine learning. Journal of Physics. 2020; 1684:012062. Available from: 10.1088/1742-6596/1684/1/012062

44. Le TM, Vo TM, Pham TT, Dao SVT. A novel Wrapper–Based feature selection for early diabetes prediction enhanced with a metaheuristic. IEEE Access. 2021; 9: 7869–84. Available from: 10.1109/access.2020.3047942

45. Julius AO, Ayokunle AO, Ibrahim FO. Early diabetic risk prediction using machine learning classification techniques. International Journal of Innovative Science and Research Technology. 2021; 6(9): 502.

46. Hassan AS, Malaserene I, Leema AA. Diabetes Mellitus Prediction using Classification Techniques. International Journal of Innovative Technology and Exploring Engineering. 2020; 9(5): 2080–4. Available from: 10.35940/ijitee.e2692.039520

47. Birjais R, Mourya AK, Chauhan R, Kaur H. Prediction and diagnosis of future diabetes risk: a machine learning approach. SN Applied Sciences. 2019; 1: 1–8. Available from: 10.1007/s42452-019-1117-9

48. Phongying M, Hiriote S. Diabetes classification using machine learning techniques. Computation (Basel). 2023; 11(5): 96. Available from: 10.3390/computation11050096

49. Maniruzzaman Md, Rahman MdJ, Ahammed B, Abedin MdM. Classification and prediction of diabetes disease using machine learning paradigm. Health Information Science and Systems. 2020; 8(1). Available from: 10.1007/s13755-019-0095-z

